# Beyond the Reading Room: Integrating Radiology into a Longitudinal Integrated Framework

**DOI:** 10.1101/2023.01.29.23285166

**Authors:** Denisse Cristina Porras Fimbres, Emily Hatheway Marshall, Alexis Musick, Robert French, Jonathan Martin, Poonam Sharma

**Affiliations:** Medical student, Duke University School of Medicine, Durham, NC, USA; Department of Radiology, Division of Musculoskeletal Ultrasound, Duke University School of Medicine, Durham, NC, USA; Department of Radiology, Division of Interventional Radiology, Duke University School of Medicine, Durham, NC, USA; Department of Medicine, Division of General Internal Medicine, Duke University School of Medicine, Durham, NC, USA

**Keywords:** Longitudinal Integrated Clerkships, integrated radiology curriculum, systems-based education, patient-centered radiology, blended LIC

## Abstract

**Introduction:** Although longitudinal integrated clerkship models have a growing evidence base and prevalence, the integration of radiology in such models is rarely explored. Institutions that have included radiology in longitudinal programs favor block rotation-style models, which do not capture longitudinal clinical care relationships with patients. We aimed to incorporate radiology learning objectives in a patient-centered and systems-based approach in a Longitudinal Integrated Clerkship pilot radiology program.

**Methods:** Medical students participating in the Longitudinal Integrated Clerkship model (N=9) chose imaging encounters from their patient panel or longitudinal clinical experiences and coordinated individual reviews with radiology faculty. 25 unique imaging encounters were required for this pilot program. This replaced discrete, asynchronous half-day experiences across radiology subspecialties that did not leverage patients that Longitudinal Integrated Clerkship students saw in clinic. Post-encounter and end-of-clerkship surveys were distributed to pilot students, and feedback surveys were distributed to faculty. Results were analyzed using descriptive statistics; end-of-clerkship data was compared to three pre-pilot academic years.

**Results:** Longitudinal Integrated Clerkship students logged 237 unique radiology encounters with a maximum individual encounter log of 33 encounters and a mean of 26 encounters. All students were exposed to at least three of the most common imaging modalities: radiographs, computed tomography scans, and magnetic resonance imaging examinations. According to faculty evaluations, 100% of students were prepared for each imaging discussion and were above (39%) or at (61%) the level expected of their training. All students reported an increase in their skills and knowledge related to imaging or procedure indications.

**Discussion:** Leveraging the Longitudinal Integrated Clerkship framework to include radiology experiences was feasible at our institution, and student and faculty survey responses suggest that students meet their faculty expectations and that the pilot provided students with improved radiology knowledge and skills.

## Introduction

Traditionally, radiology has been taught to medical students using a combination of preclinical education (e.g., didactic lectures, case-based learning) and clinical clerkships or electives.^[1–6]^ In the United States, 16% of medical schools require a dedicated radiology clerkship, the vast majority of which are standalone block rotations (BRs).^[1,2,5–8]^

Despite the increasing prevalence of medical imaging in patient care and the long-held perception of radiology as an important curricular component, there are no uniform guidelines about how or when to teach radiology to medical students.^[3,9–12]^ To help standardize radiology education, several organizations have developed radiology curricula outlining core topics.^[13,14]^ For example, the Alliance of Medical Student Educators in Radiology (AMSER), within the Association of University Radiologists (AUR), developed a national medical student curriculum that reviews technical aspects, normal anatomy, pathology, procedures, and imaging algorithms across numerous organ-based systems.^[13]^ AUR also released an additional set of sample learning objectives to be included in a curriculum.^[13]^ However, there is no robust direction on how to best deliver this education or ensure that the learning objectives are met.

While some institutions opt for traditional BRs, these condensed models miss the opportunity to better capture longitudinal clinical and professional relationships and illustrate systems-based care, prompting the increasing use of longitudinal integrated clerkships (LICs) across specialties. In contrast to sequential, time-limited BRs, LICs use a continuity-based model where students participate in the comprehensive care of a patient panel under the same preceptors, achieving clinical competencies of multiple disciplines simultaneously.^[15]^ For example, a traditional OB/GYN BR student may spend two consecutive weeks working with a variety of preceptors in an outpatient clinic; a longitudinal model may instead have students spend one half-day with the same preceptor in clinic for six months, with students following their patients into the hospital or operating rooms as needed.^[15]^ Patient and preceptor continuity allows for education tailored to the student over time, facilitating enhanced student satisfaction with faculty teaching compared to BR students.^[15–20]^ Importantly, LIC students score similarly on clinical evaluations and standardized exams when compared to their BR counterparts, suggesting that learning outcomes can withstand structural curricular changes.^[18–20]^

While LIC programs intend to meet diverse core competencies simultaneously, radiology has not been as readily incorporated into longitudinal curricula. Although several recent publications discuss the incorporation of distributed radiology content into BR models, such as online radiology cases that correspond to BR content or protected time for separate radiology didactics,^[21–28]^ studies exploring the integration of radiology with longitudinal clinical experiences are scarce. Where published, LIC programs that incorporate radiology tend to do so in a more structured fashion, including introductory didactic sessions and regular radiology rounds.^[17,29]^ Radiology-related outcomes measures in these models mirror the medical literature, with LIC students scoring minimally lower on final exams with no significant changes in other evaluation methods (e.g., Objective Structured Clinical Examination cases).^[29]^ However, these existing curricular models do not necessarily leverage the longitudinal faculty relationships and expansive patient panels intrinsic to LIC programs.

Given the increasing prevalence of LICs and the continued importance of teaching medical imaging, there is a need for additional models describing how radiology can be incorporated into a LIC. With this gap in mind, we sought to explore the feasibility of integrating a novel radiology curriculum into an existing LIC framework using a unique patient panel-centered approach. We also aimed to evaluate student satisfaction with this model and if this program provided diverse radiologic exposures.

*A subset of data was presented virtually at the Consortium of Longitudinal Integrated Clerkships (CLIC) annual conference, October 10-13, 2021. A presentation of this work will be presented at the AAMC: Learn, Serve, Lead, November 11-15, 2022, and an abstract of this work (substantially different) will be published in Academic Medicine RIME abstract supplement*.

## Materials and Methods

### Context

Duke University School of Medicine is a medical school associated with a large tertiary care center in the southeastern United States. Students participate in the clerkship year during their second academic year. There are BR and LIC models available for medical students’ clerkship years. To participate in the LIC curriculum, students complete a formal application and interview process. All Duke medical students are required to complete a radiology clerkship, regardless of clerkship modality.

Prior to the 2020-2021 academic year (AY), the LIC radiology clerkship was composed of 16 half-days in various radiology specialties throughout the six-month longitudinal outpatient portion of the year. Both the curriculum and the cases discussed with radiologists were not specific to LIC students’ patient panels in this model. Instead, students were assigned faculty contacts on specific half-days throughout the year across different imaging subspecialties. All BR students participate in a 4-week traditional BR in radiology.

### Program Design

In the pilot program (2020-2021 AY) LIC students identified patients during their clinical experiences (i.e., clinics, urgent care, emergency department, or inpatient immersion) who would be receiving radiology examinations or procedures. Students were encouraged to accompany the patient to the radiology studies/procedures. Students were required to meet individually with a radiologist to review the clinical indications and imaging findings of the examination for a minimum of 25 patient cases throughout the clerkship year. There were no specific guidelines to the breadth of cases or a requirement for the variety of imaging modalities during this pilot program. Students were provided the radiology didactic materials utilized by the BR students. They were expected to asynchronously review these materials, which were composed of handouts, case reviews, and recorded lectures.

Radiology faculty (17) were identified in each imaging subspecialty and a “point of contact” list was provided to the LIC students by the clerkship directors. Faculty who had participated in medical student teaching in the past were invited to participate. There were no additional incentives for faculty members to become part of the pilot program. The radiologists were provided faculty development on both LIC and the pilot program.

Students emailed radiologists from the “point of contact” list and met individually with the radiology attending physicians to discuss their case(s) relevant to the radiologist’s field of expertise. The time spent discussing cases was determined by student and faculty availability and was individual to each student experience. Students could meet with multiple radiologists in the same week to discuss cases if their schedule permitted. Students met one-on-one with these faculty members and were required to present their patient cases to the radiologists (orally or using picture archiving and communication system [PACS] stations) and describe their reading of the image. The radiologists would then provide education on how to systematically read the imaging modality and allow for questions based upon the specific case or the imaging type in general. The exact structure of these encounters was dependent on student engagement and faculty teaching style.

Students in the pilot cohort were graded on a satisfactory and unsatisfactory scale and assessed based on faculty feedback and performance during each encounter. Prior LIC cohorts did not receive a formal grade for the clerkship experience.

### Analysis

LIC students were required to choose 10 faculty members that they had worked with to provide written formative feedback during the pilot program. This evaluation was a 10-question survey distributed through the Qualtrics platform (Appendix A). The survey asked about patient demographic data and imaging exam details but focused primarily on student preparedness for the encounter, what the student did well, and how the student could improve in subsequent encounters.

Student participation was tracked via an 11-question Qualtrics survey completed following each meeting with a radiologist (Appendix B). Survey questions included demographic information and presenting symptoms for the patient as well as the imaging modality and the diagnostic impression after speaking with the radiologist. The survey also asked about the appropriateness of the examination, if the exam confirmed clinical suspicions, and if it changed or directed clinical management.

LIC students were given an optional end-of-clerkship (EOC) survey as part of routine School of Medicine educational assessments to rate their experience with the radiology clerkship (Appendix C). Data from AYs 2017-2018, 2018-2019, 2019-2020 were collected. For AY 2020-2021, given the change to the new pilot program, the same survey was not collected at the end of the year; instead, a subset of questions was distributed to the LIC cohort in January 2022. One participant recused themself from the survey as they became a co-author of this paper (EHM).

All survey data was analyzed using descriptive statistics.

## Results

The nine LIC students logged 237 unique radiology encounters during the pilot program (Figure 1). Eight students reached the target of 25 radiology encounters with a mean of 26.33 encounters logged. The student who did not reach the target of 25 encounters instead logged 24 encounters. The maximum number logged by one individual was 33 encounters.

**Figure 1.**
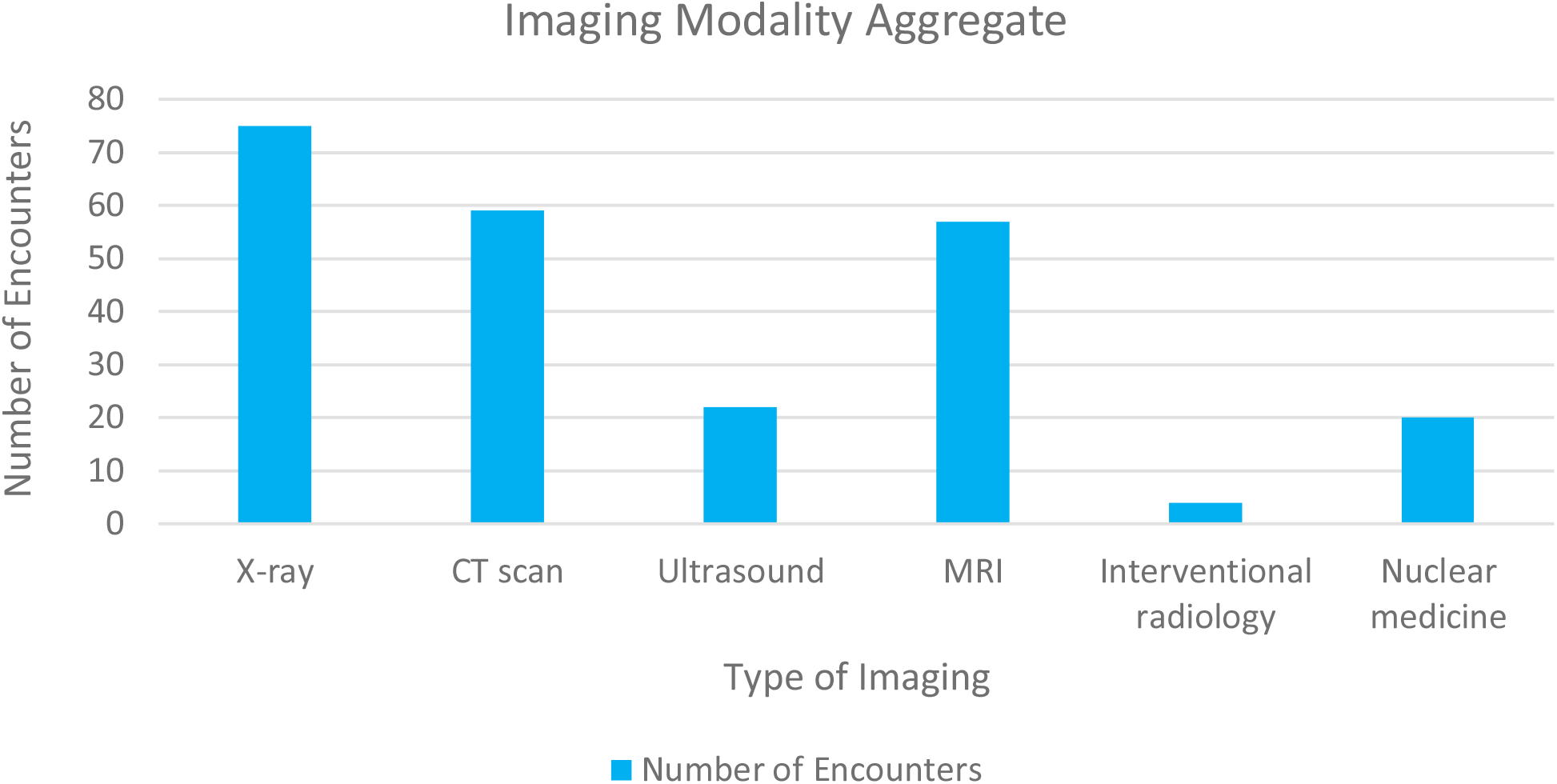
Number of unique encounters with each imaging modality for N=237 radiology encounters from all (N=9) LIC students in the program

At the student-specific level, every student had exposure to at least four unique imaging modalities. All students had at least two encounters with radiographs, magnetic resonance imaging (MRI), and computed tomography (CT) scans, and 89% (8/9) of students completed at least one ultrasound encounter. Exposure to nuclear medicine (67%) and interventional radiology (IR) (22%) was variable. Aggregate data by modality (Figure 2) and body part (Figure 3) for the pilot cohort are visualized.

**Figure 2.**
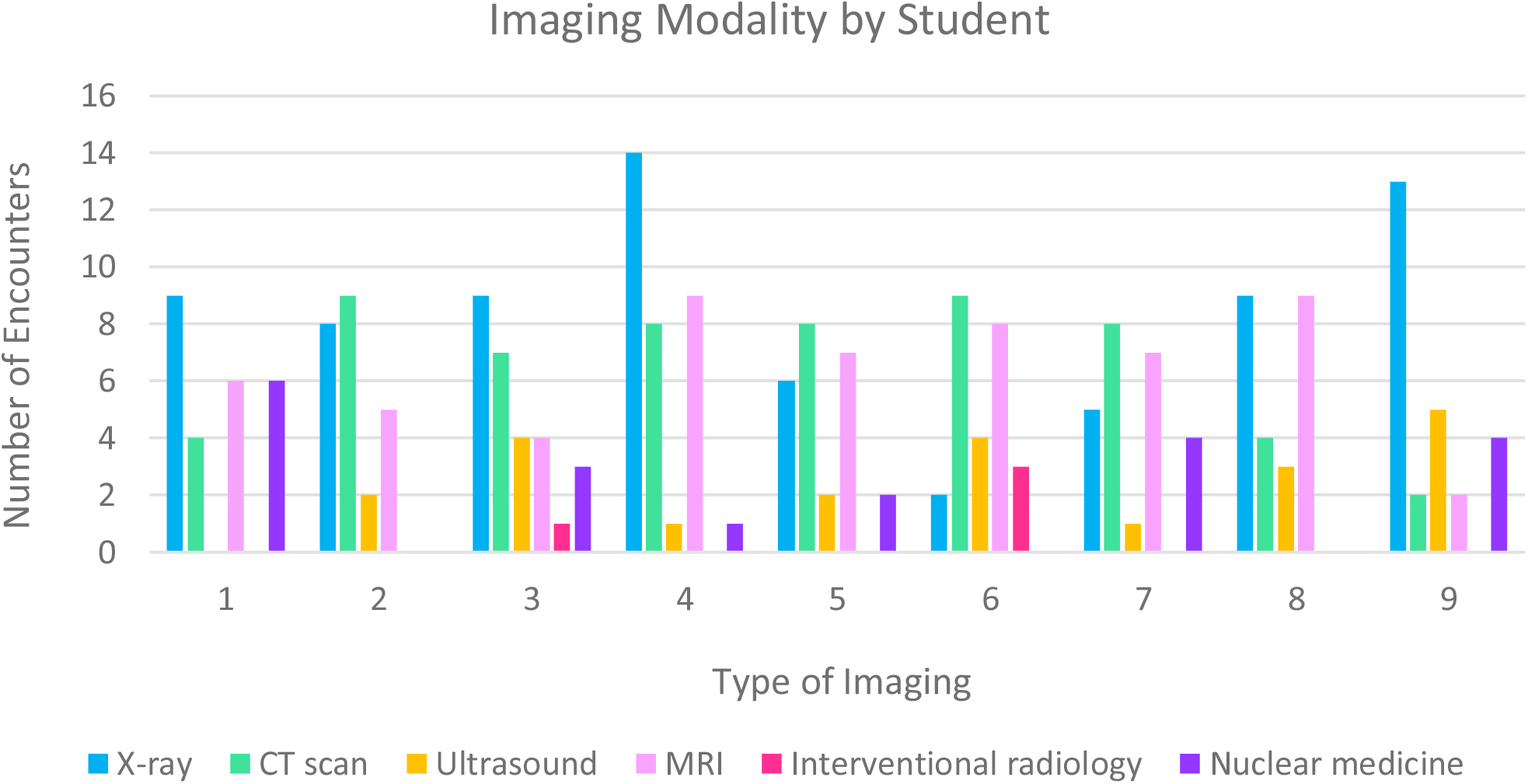
Imaging modalities documented per (N=9) students in the program

**Figure 3.**
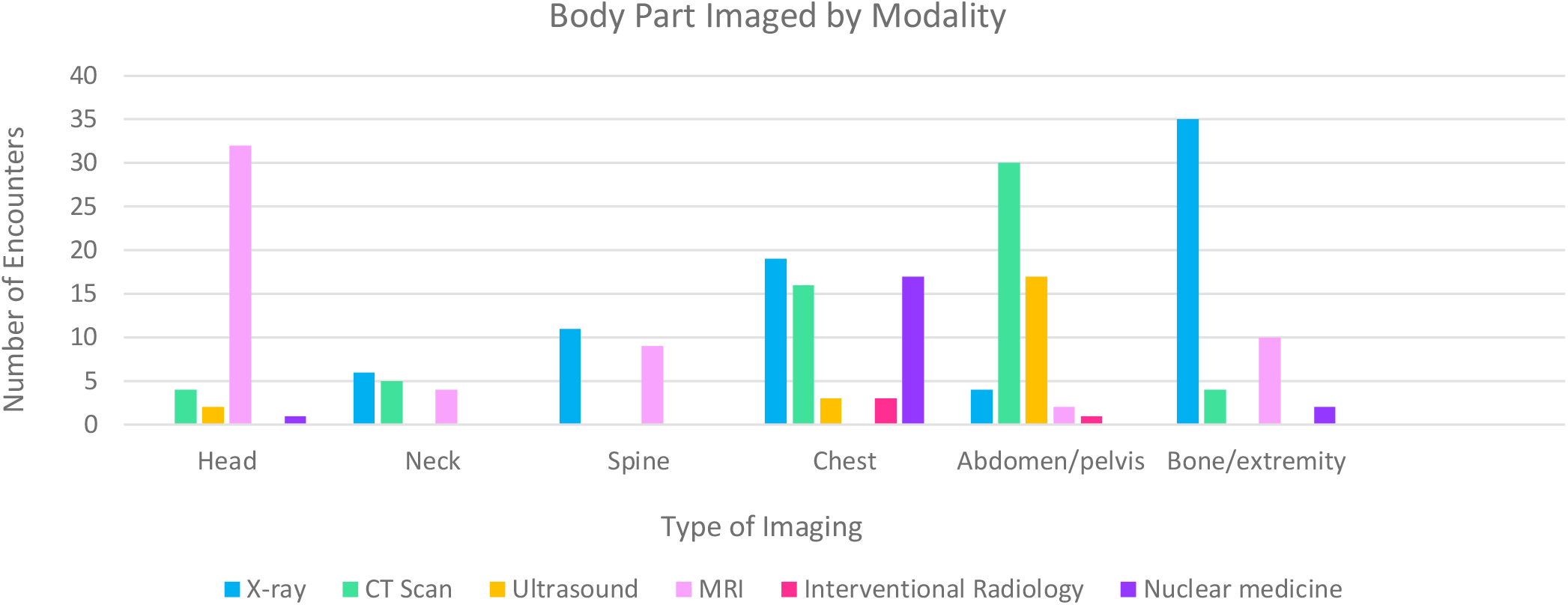
The body part imaged displayed by imaging modality for (N=237) documented radiologist encounters from all (N=9) LIC students in the program

Twelve faculty members completed a total of 84 feedback evaluations, with a mean of 9.3 pieces of feedback per LIC student. These faculty members responded that LIC students were above (39%) or at (61%) the level expected of their training, and 100% of students were prepared for the interaction with the radiologist. Of the 84 instances with narrative feedback, a good understanding of the patient and clinical scenario or the history was mentioned in 77% of the evaluations. For areas of improvement, none, NA, or did everything well, were mentioned in 48% of the evaluations. 25% of the evaluations mentioned expanding differential diagnoses as an area to improve.

In the EOC survey, most students in each academic year rated the clerkship as average or above average (Figure 4). 71% of pilot students responded that the clerkship activities almost always or regularly helped them achieve the stated goals; this is an increase, as pre-pilot cohorts’ responses to this question ranged from 25-50% (Figure 5). Across all cohort class years, all but two students (both in AY 2018-2019) reported their knowledge of the indications for imaging exams and procedures significantly increased, moderately increased, or slightly increased (Figure 6).

**Figure 4.**
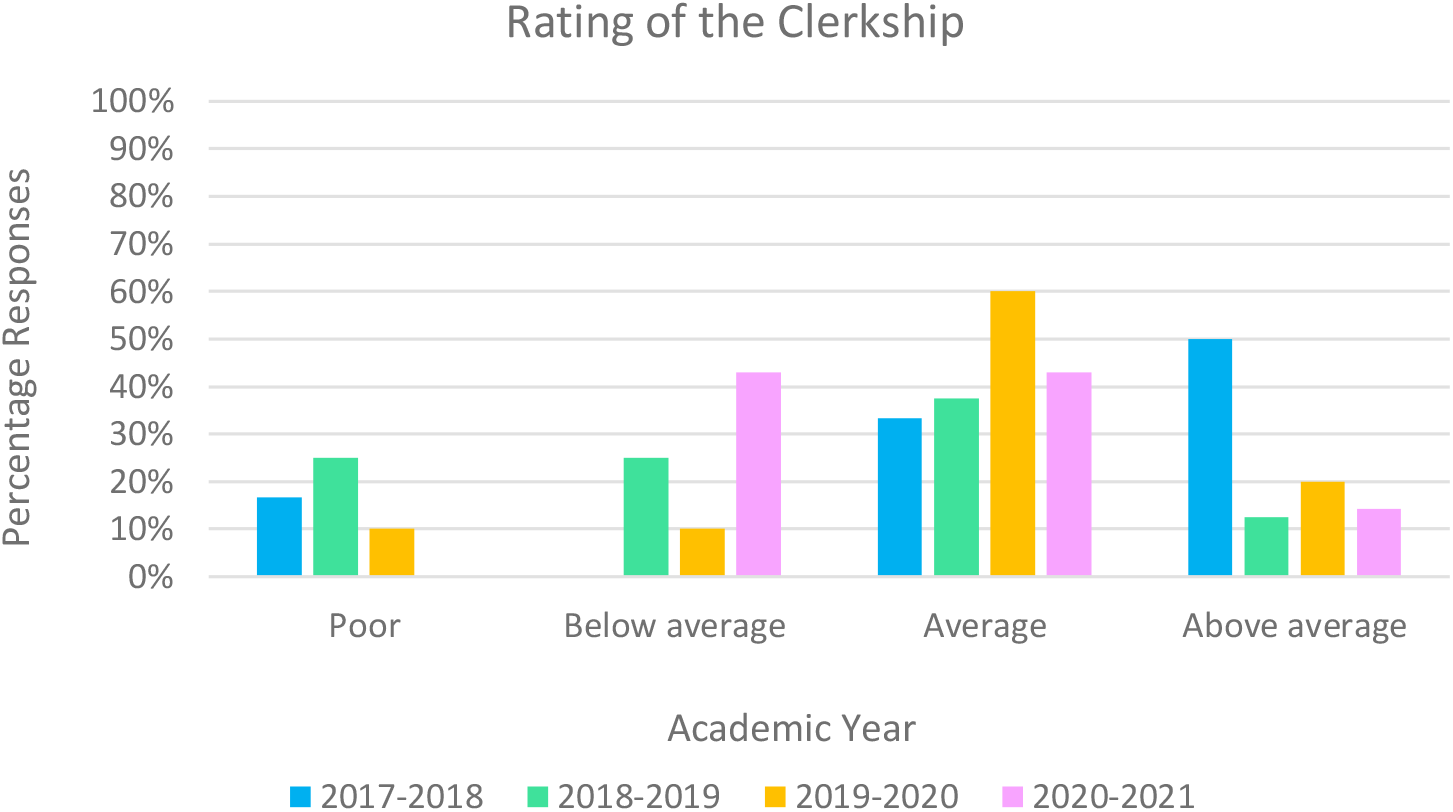
Overall rating of the Clerkship across four academic years (2017-2018, 2018-2019, 2019-2020, 2020-2021*). *Pilot cohort year

**Figure 5.**
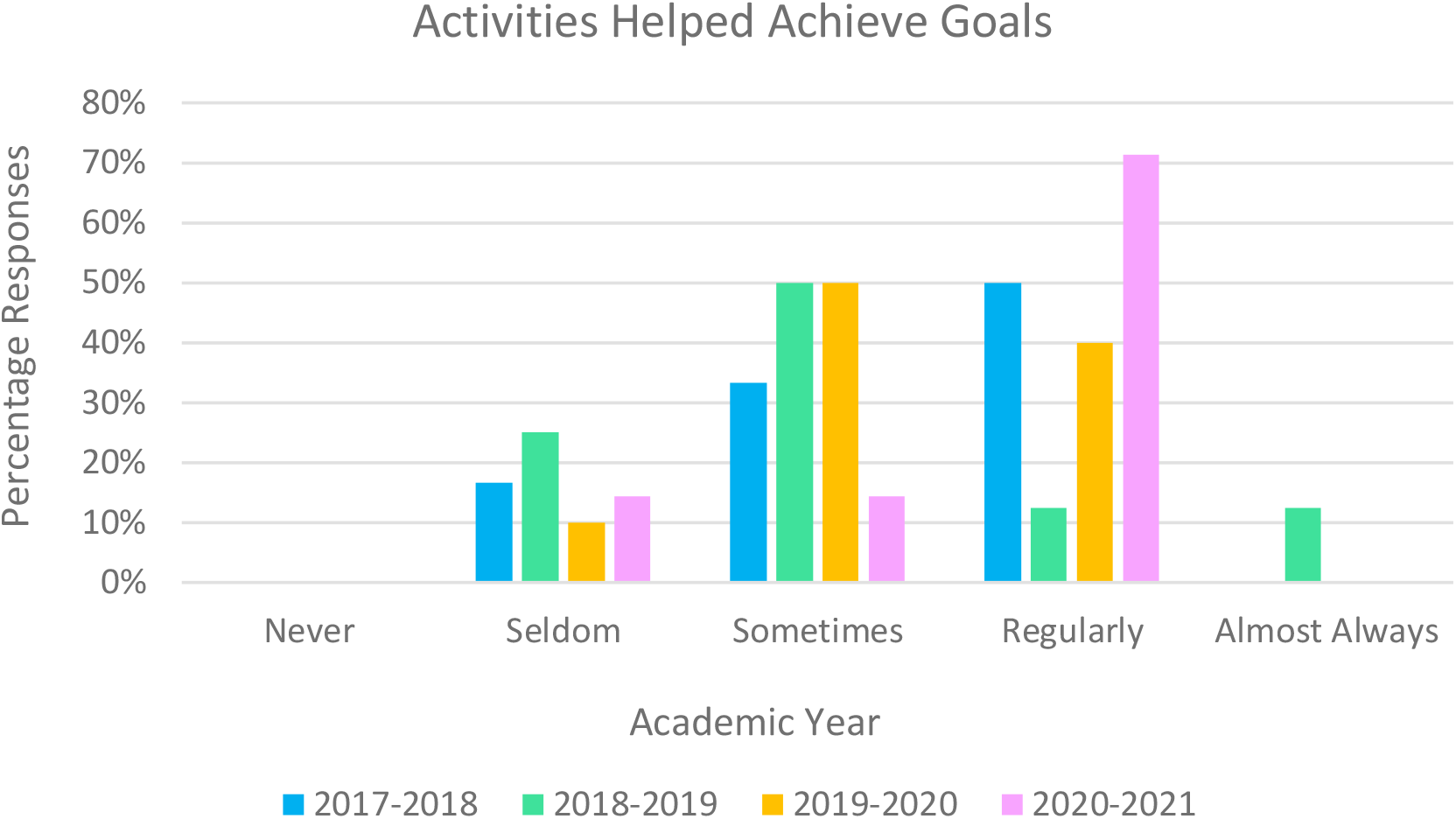
Rating of ‘clerkship activities helped achieve the stated goals’ across four academic years (2017-2018, 2018-2019, 2019-2020, 2020-2021*). *Pilot cohort year

**Figure 6.**
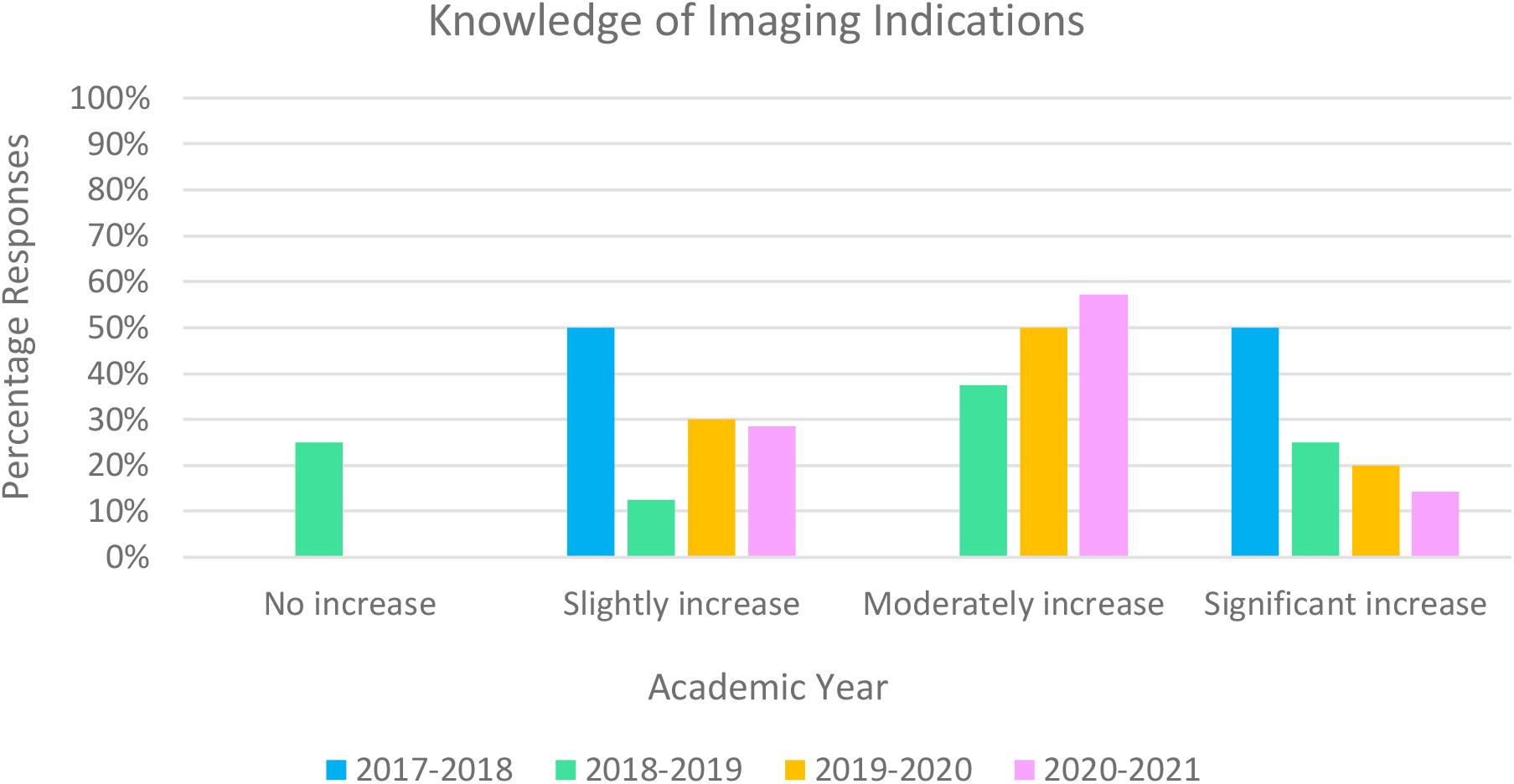
Rating of ‘knowledge of the indications for common imaging exams and procedures’ across four academic years (2017-2018, 2018-2019, 2019-2020, 2020-2021*). *Pilot cohort year

Similarly, all but one student (also in AY 2018-2019) reported their skills in image interpretation significantly increased, moderately increased, or slightly increased. 86% of pilot students thought the amount of patient care responsibility was appropriate compared to 38-60% for pre-pilot students (Figure 7). 86% of students in the pilot cohort reported the amount of supervision from preceptors was appropriate, while for students in pre-pilot cohorts it was 38-60% (Figure 8).

**Figure 7.**
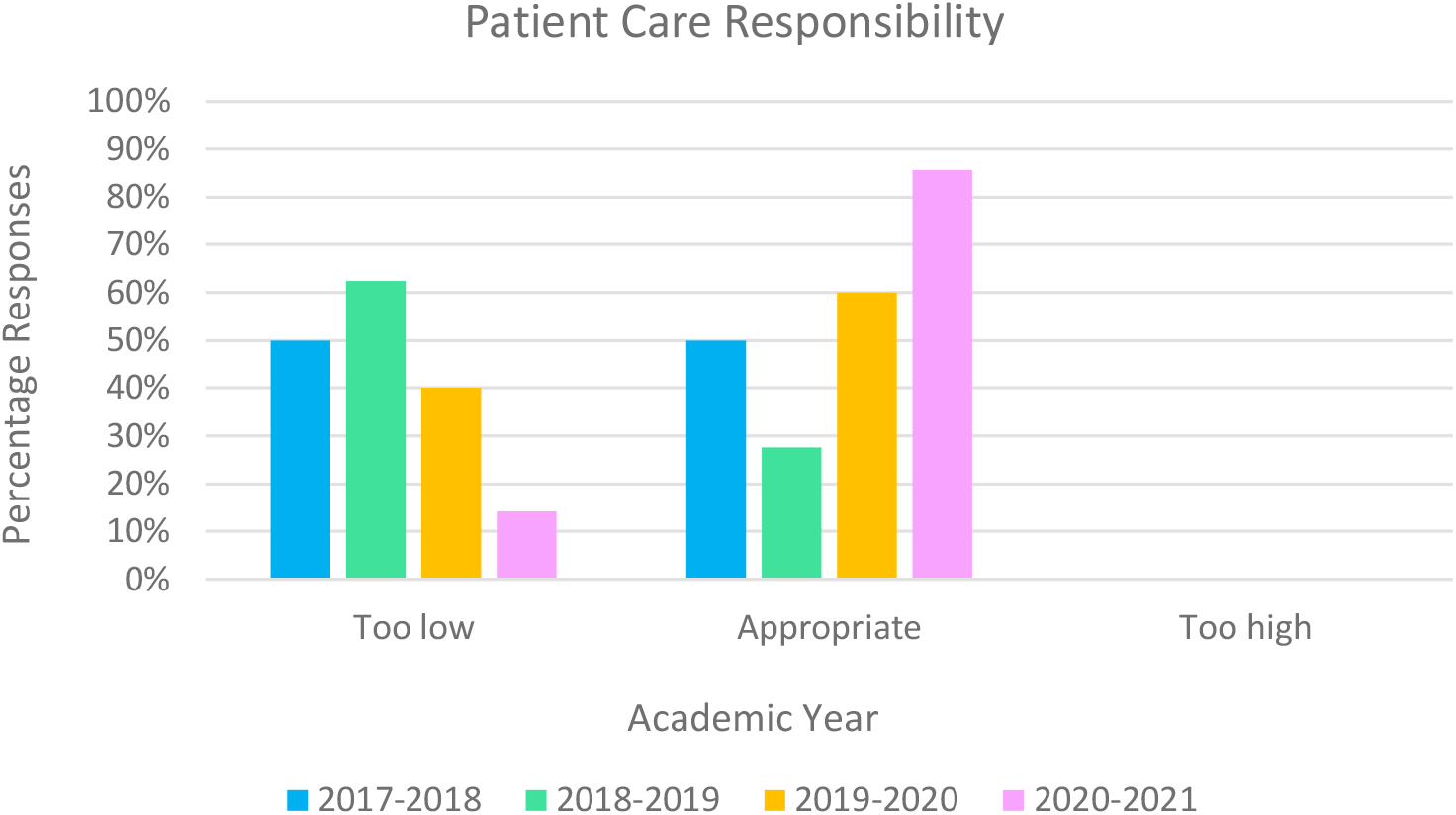
Rating of ‘Amount of patient care responsibility given’ across four academic years (2017-2018, 2018-2019, 2019-2020, 2020-2021*). *Pilot cohort year

**Figure 8.**
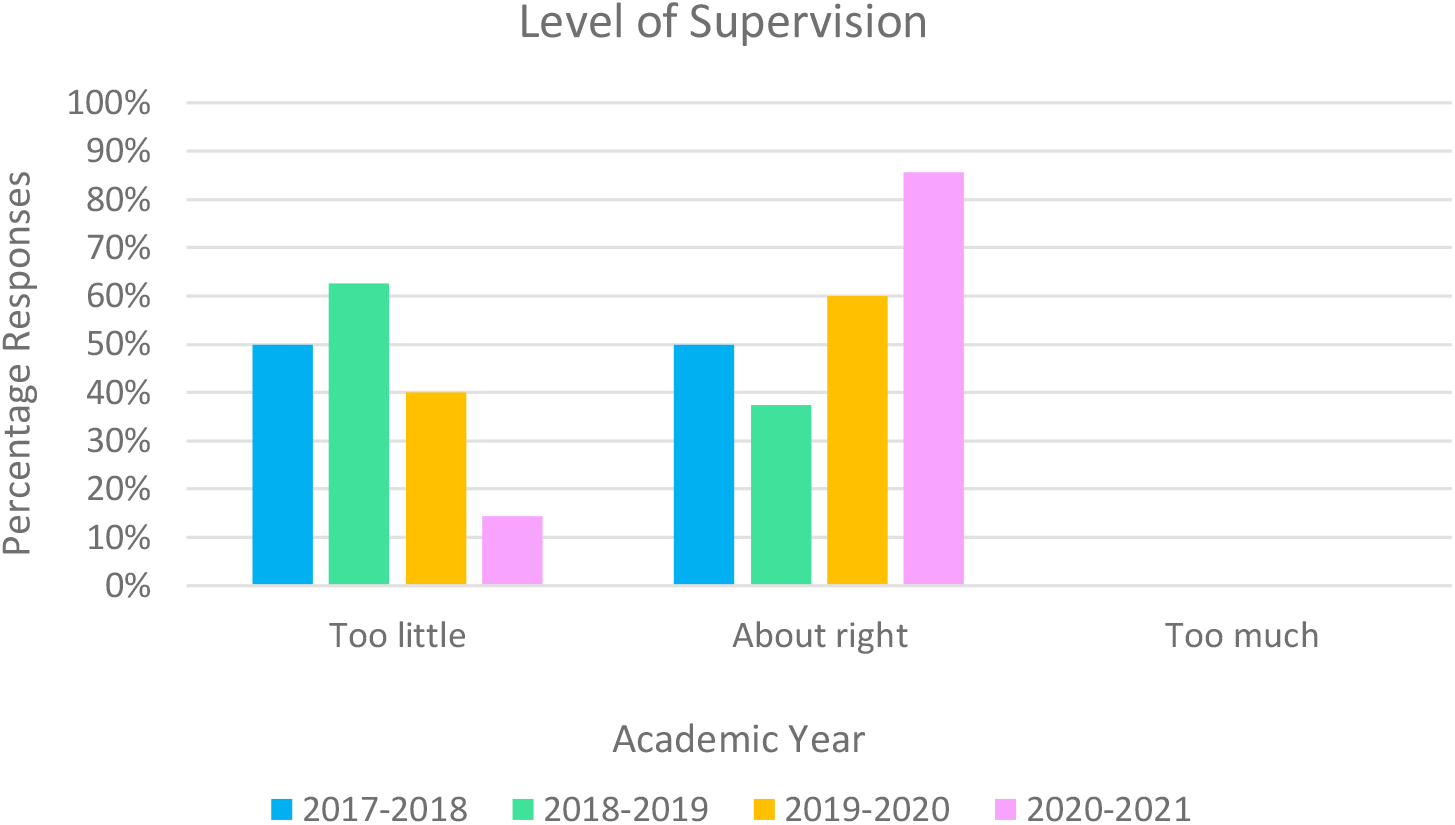
Rating of ‘level of supervision from preceptors/attendings’ across four academic years (2017-2018, 2018-2019, 2019-2020, 2020-2021*). *Pilot cohort year

## Discussion

Our study suggests that it is feasible to incorporate radiology education into the structure of an LIC at our institution. We found that students reported increased knowledge and improved skills in imaging interpretation at levels comparable to prior cohorts. Pilot students also reported that the clerkship structure helped them achieve the stated goals at a rate comparable to pre-pilot years. Importantly, students in the pilot more frequently noted patient care responsibilities and preceptor supervision as being at an appropriate level. Additionally, all faculty evaluators reported that students were prepared for the interactions and were at or above their expected level of training. These results attest to the potential of an integrated LIC radiology clerkship to provide instruction that is comparable to traditional BRs.

These findings show that students had diverse imaging educational experiences at an individual level and had at least some varied exposure to commonly used modalities. Since there were no imaging-specific requirements, this variation most likely reflects the diverse patient population followed by the LIC students. This variability could also be explained by the diverse subspecialty clinics the students attend. In future cohorts, it will be important to investigate student empowerment given the increase in perceived patient care responsibility reported in the survey results. As the program continues and expands, further exploration in this area may elucidate trends among the type and number of patients the LIC students follow.

Students reported improved knowledge about the indications for imaging on the EOC survey, which is an essential skill of ordering clinicians of the future in accordance with suggested AUR and AMSER learning objectives. By leveraging LIC patient panels in this pilot, students were able to see the spectrum of care in a consultative role with radiologists. Our results show that using the patient-centered framework outlined in the pilot curriculum is a feasible model that addresses objectives outlined by governing bodies in a novel way.

While exposure to some fields within radiology such as IR was low, students generally had a wide breadth of experiences in other areas. The low exposure observed could be due to student preference or the patient population each student followed. Given IR is procedure-based and cases are usually during the day, except for add-ons or emergency cases, perhaps students had difficulty attending from a logistical standpoint. Furthermore, IR cases are live, which precludes students from reviewing imaging at a more flexible time unlike other subspecialties such as ultrasound, CT or MRI. Moving forward, it will be important to continually assess the diversity of imaging experiences in future cohorts.

There are limitations to this study. The pilot program took place at a single institution with a small sample size. The sample size wholly represented the target population, however. The radiology educational experiences for the pre-pilot and pilot cohorts were, by design, fragmented across the year and may not have been accurately recalled and reflected upon in the EOC survey. A lower number of pilot students rated the clerkship as average or above average (57%) relative to the prior academic year, but this is not necessarily a reflection of the clerkship itself; these ratings historically fluctuate (50-83% in pre-pilot years) and may reflect other variables that were not controlled for. The internal comparison metrics, including accomplishing the required encounters, achieving stated goals, and improving skills in image interpretation, were higher among the pilot students, supporting the success of this novel program. There were no inferential statistics performed and findings reported were observational, which limits the identification of a cause-effect relationship. Additionally, there are drawbacks to LIC models. Longitudinal clerkships are less structured than traditional rotations which for some students may be challenging while for others it is beneficial.^[30]^ There may also be added burden on faculty mentors to find a time in their clinical schedule to meet with students and ensure patient care is not compromised.^[30]^

## Conclusion

This LIC pilot radiology study suggests that incorporating a radiology curriculum into an LIC by leveraging established LIC patient relationships is effective and feasible. This is a novel model for medical student radiology educational delivery focused on a patient-centered approach. It is a promising model for institutions that seek to incorporate a robust and patient-centered radiology education into a LIC. This model could also be incorporated into a more traditional BR framework.

## Data Availability

All data produced in the present study are available upon reasonable request to the authors

## Acknowledgements

*The authors wish to acknowledge Dr Deborah Engle, for assisting in the development of assessment strategies of longitudinal integrated clerkships utilized in this work*.

### Appendix A. Faculty Feedback Form

|  |
| --- |
| 1. Select a medical student you are evaluating and answer the following questions |
| 2. On what services did you interact with this student (select all that apply)? |
| a. Interventional Radiology |
| b. Musculoskeletal Imaging |
| c. Abdominal Imaging |
| d. Nuclear Medicine |
| e. Cardiothoracic Imaging |
| f. Breast Imaging |
| g. Neuroradiology |
| h. Pediatric Radiology |
| 3. Imaging modality: |
| a. X-rays |
| b. Ultrasound |
| c. Computed tomography |
| d. Magnetic resonance imaging |
| e. Nuclear Medicine |
| f. Procedure (any) |
| g. PET/CT |
| h. Mammography |
| 4. Body area imaged: |
| a. Chest |
| b. Abdomen/Pelvis |
| c. Bone/Extremity |
| d. Head |
| e. Neck/Spine |
| f. Breast |
| 5. Presenting symptom |
| 6. Was the student well prepared for this interaction? |
| a. Yes |
| b. No |
| 7. Was the interaction at the expected level of training for the student? |
| a. Above expected level of training |
| b. At expected level of training |
| c. Below expected level of training |
| 8. What did the student do particularly well during the interaction? |
| 9. What are areas for improvement in subsequent encounters? |
| 10. Please provide your name. By typing your name, you certify that you have not provided direct health care services to this student and that you will uphold the Duke Code of Professional Conduct (this is an LCME requirement). |

### Appendix B Student Form-Post-Radiologist Interaction

|  |
| --- |
| 1. Student name |
| 2. Patient age |
| 3. Patient reported gender |
| 4. Presenting symptoms |
| 5. Imaging modality |
| 6. Body part imaged |
| 7. Impression of imaging exam |
| 8. Name of reviewing radiologist |
| 9. Appropriateness of examination |
| 10. Did the exam confirm the clinical suspicion? |
| 11. Did the exam change or direct clinical management? |

### Appendix C End-of-Year Survey

|  |
| --- |
| 1. All things considered, what is your overall rating of this clerkship? |
| a. Poor |
| b. Below average |
| c. Average |
| d. Above average |
| 2. Clerkship goals and objectives were clearly stated in the orientation materials and sub-rotation packets: |
| a. No |
| b. Somewhat |
| c. Yes |
| 3. Clerkship activities helped me achieve the stated goals: |
| a. Never |
| b. Seldom |
| c. Sometimes |
| d. Regularly |
| e. Almost always |
| 4. Clerkship grading and the elemental components of the grade were clear: |
| a. Never |
| b. Seldom |
| c. Sometimes |
| d. Regularly |
| e. Almost always |
| 5. Feedback from preceptors/attendings were constructive: |
| a. Never |
| b. Seldom |
| c. Sometimes |
| d. Regularly |
| e. Almost always |
| 6. The amount of patient care responsibility I was given was: |
| a. Too low |
| b. Appropriate |
| c. Too high |
| 7. The level of supervision from preceptors/attendings was: |
| a. Too little |
| b. About right |
| c. Too much |
| 8. Please rate your satisfaction with the amount of formative feedback in this clerkship: |
| a. Very dissatisfied |
| b. Dissatisfied |
| c. Satisfied |
| d. Very satisfied |
| 9. Please rate your satisfaction with the quality of formative feedback in this clerkship: |
| a. Very dissatisfied |
| b. Dissatisfied |
| c. Satisfied |
| d. Very satisfied |
| 10. This clerkship helped me learn how to better utilize imaging to evaluate common medical problems: |
| a. Strongly disagree |
| b. Mildly disagree |
| c. Mildly agree |
| d. Strongly agree |
| 11. In general, the faculty on this rotation seemed invested in medical student education: |
| a. Strongly disagree |
| b. Mildly disagree |
| c. Mildly agree |
| d. Strongly agree |
| 12. The volume of patients was: |
| a. Too low |
| b. Appropriate |
| c. Too high |
| 13. Do you believe that what you learned and experienced during your radiology clerkship will help you be a better doctor and help you better utilize imaging in patient care, regardless of your chosen specialty? |
| a. Strongly disagree |
| b. Disagree |
| c. Neutral |
| d. Agree |
| e. Strongly agree |
| 14. How much did your SKILL LEVEL increase in the following areas as a result of the clerkship? |
| a. Skill in using the electronic medical record and PACS use for History taking |
| i. No increase |
| ii. Slightly increased |
| iii. Moderately increased |
| iv. Significantly increased |
| b. Knowledge of the pros and cons of common imaging exams and procedures enabling better image utilization in patient care: |
| i. No increase |
| ii. Slightly increased |
| iii. Moderately increased |
| iv. Significantly increased |
| c. Imaging interpretation skills |
| i. No increase |
| ii. Slightly increased |
| iii. Moderately increased |
| iv. Significantly increased |
| d. Differential diagnosis skills |
| i. No increase |
| ii. Slightly increased |
| iii. Moderately increased |
| iv. Significantly increased |
| e. Knowledge of the indications for common imaging exams and procedures in radiology |
| i. No increase |
| ii. Slightly increased |
| iii. Moderately increased |
| iv. Significantly increased |
| f. Knowledge of the pros and cons of common imaging exams and procedures enabling better image utilization in patient care |
| i. No increase |
| ii. Slightly increased |
| iii. Moderately increased |
| iv. Significantly increased |
| g. Presentation skills |
| i. No increase |
| ii. Slightly increased |
| iii. Moderately increased |
| iv. Significantly increased |
| 15. For the lecture series, please comment on any especially good lectures |
| 16. For the lecture series, please comment on any especially poor lectures |
| 17. Please list (and explain why) any faculty who were particularly effective teachers |
| 18. Please list (and explain why) any faculty who were NOT effective teachers |
| 19. What did you consider to be the most valuable aspects of this clerkship? |
| 20. What are areas for improvement in this clerkship? |

## References

1. Barlev DM, Lautin EM, Amis ES, Lerner ME. A survey of radiology clerkships at teaching hospitals in the United States. Invest Radiol. 1994;29(1):105–108. doi:10.1097/00004424-199401000-00020

2. Chew C, Cannon P, O’Dwyer PJ. Radiology for medical students (1925–2018): an overview. BJR Open. 2020;2(1). doi:10.1259/bjro.20190050

3. Dmytriw AA, Mok PS, Gorelik N, Kavanaugh J, Brown P. Radiology in the Undergraduate Medical Curriculum: Too Little, Too Late? Med Sci Educ. 2015;25(3):223–227. doi:10.1007/s40670-015-0130-x

4. Kourdioukova EV, Valcke M, Derese A, Verstraete KL. Analysis of radiology education in undergraduate medical doctors training in Europe. Eur J Radiol. 2011;78(3):309–318. doi:10.1016/j.ejrad.2010.08.026

5. Lee H, Kim DH, Hong PP. Radiology Clerkship Requirements in Canada and the United States: Current State and Impact on Residency Application. J Am Coll Radiol. 2020;17(4):515–522. doi:10.1016/j.jacr.2019.11.026

6. Straus CM, Webb EM, Kondo KL, et al. Medical Student Radiology Education: Summary and Recommendations From a National Survey of Medical School and Radiology Department Leadership. J Am Coll Radiol. 2014;11(6):606–610. doi:10.1016/j.jacr.2014.01.012

7. LCME Annual Medical School Questionnaire Part II, 2011-2012 through 2019-2020 on AAMC. Clerkship Requirements by Discipline. AAMC. Accessed January 21, 2022. https://www.aamc.org/data-reports/curriculum-reports/interactive-data/clerkship-requirements-discipline

8. Slanetz PJ, Naeger DM, Avery LL, Deitte LA. Mixed Practice in Radiology Education— Has the Time Come? J Am Coll Radiol. 2020;17(7):976–978. doi:10.1016/j.jacr.2019.12.005

9. du Cret RP, Weinberg EJ, Sellers TA, Seybolt LM, Kuni CC, Thompson WM. Role of radiology in medical education: perspective of nonradiologists. Acad Radiol. 1994;1(1):70–74. doi:10.1016/s1076-6332(05)80789-5

10. Poot JD, Hartman MS, Daffner RH. Understanding the US Medical School Requirements and Medical Students’ Attitudes about Radiology Rotations. Acad Radiol. 2012;19(3):369–373. doi:10.1016/j.acra.2011.11.005

11. Smith-Bindman R, Kwan ML, Marlow EC, et al. Trends in Use of Medical Imaging in US Health Care Systems and in Ontario, Canada, 2000-2016. JAMA. 2019;322(9):843–856. doi:10.1001/jama.2019.11456

12. Subramaniam RM, Beckley V, Chan M, Chou T, Scally P. Radiology curriculum topics for medical students: students’ perspectives. Acad Radiol. 2006;13(7):880–884. doi:10.1016/j.acra.2006.02.034

13. Association of University Radiologists. AMSER Curriculum, Competencies, and Learning Objectives. AUR. Published 2021. Accessed December 8, 2021. https://www.aur.org/affinity-groups/amser/curriculum

14. European Society of Radiology (ESR). Undergraduate education in radiology. A white paper by the European Society of Radiology. Insights Imaging. 2011;2(4):363–374. doi:10.1007/s13244-011-0104-5

15. Worley P, Couper I, Strasser R, et al. A typology of longitudinal integrated clerkships. Med Educ. 2016;50(9):922–932. doi:10.1111/medu.13084

16. Hirsh D, Gaufberg E, Ogur B, et al. Educational outcomes of the Harvard Medical School-Cambridge integrated clerkship: a way forward for medical education. Acad Med J Assoc Am Med Coll. 2012;87(5):643–650. doi:10.1097/ACM.0b013e31824d9821

17. Ogur B, Hirsh D, Krupat E, Bor D. The Harvard Medical School-Cambridge Integrated Clerkship: An Innovative Model of Clinical Education. Acad Med. 2007;82(4):397–404. doi:10.1097/ACM.0b013e31803338f0

18. Poncelet A, Bokser S, Calton B, et al. Development of a longitudinal integrated clerkship at an academic medical center. Med Educ Online. 2011;16. doi:10.3402/meo.v16i0.5939

19. Teherani A, Irby DM, Loeser H. Outcomes of different clerkship models: longitudinal integrated, hybrid, and block. Acad Med J Assoc Am Med Coll. 2013;88(1):35–43. doi:10.1097/ACM.0b013e318276ca9b

20. Latessa RA, Swendiman RA, Parlier AB, Galvin SL, Hirsh DA. Graduates’ Perceptions of Learning Affordances in Longitudinal Integrated Clerkships: A Dual-Institution, Mixed-Methods Study. Acad Med J Assoc Am Med Coll. 2017;92(9):1313–1319. doi:10.1097/ACM.0000000000001621

21. Chew FS, Relyea-Chew A. Distributed Web-supported radiology clerkship for the required clinical clerkship year of medical school: development, implementation, and evaluation. Acad Radiol. 2002;9(6):713–720. doi:10.1016/s1076-6332(03)80317-3

22. Chew FS. Distributed Radiology Clerkship for the Core Clinical Year of Medical School. Acad Med. 2002;77(11):1162–1163.

23. Chorney ET, Lewis PJ. Integrating a Radiology Curriculum Into Clinical Clerkships Using Case Oriented Radiology Education. J Am Coll Radiol. 2011;8(1):58-64.e4. doi:10.1016/j.jacr.2010.08.018

24. Collins J, Dotti SL, Albanese MA. Teaching radiology to medical students: an integrated approach. Acad Radiol. 2002;9(9):1046–1053. doi:10.1016/s1076-6332(03)80481-6

25. Di Salvo DN, Clarke PD, Cho CH, Alexander EK. Redesign and Implementation of the Radiology Clerkship: From Traditional to Longitudinal and Integrative. J Am Coll Radiol. 2014;11(4):413–420. doi:10.1016/j.jacr.2013.05.018

26. Lim-Dunham JE, Ensminger DC, McNulty JA, Hoyt AE, Chandrasekhar AJ. A Vertically Integrated Online Radiology Curriculum Developed as a Cognitive Apprenticeship:: Impact on Student Performance and Learning. Acad Radiol. 2016;23(2):252–261. doi:10.1016/j.acra.2015.09.018

27. Lowitt NR. Assessment of an Integrated Curriculum in Radiology. Acad Med. 2002;77(9):933.

28. Tigges S, Lewis PJ, McNulty NJ, Mullins ME. Medical Student Performance After a Vertically Integrated Radiology Clerkship. J Am Coll Radiol. 2016;13(1):67–71. doi:10.1016/j.jacr.2015.07.019

29. Shaffer K, Ng JM, Hirsh DA. An integrated model for radiology education: development of a year-long curriculum in imaging with focus on ambulatory and multidisciplinary medicine. Acad Radiol. 2009;16(10):1292–1301. doi:10.1016/j.acra.2009.06.002

30. Hense H, Harst L, Küster D, Walther F, Schmitt J. Implementing longitudinal integrated curricula: Systematic review of barriers and facilitators. Med Educ. 2021;55(5):558–573. doi:10.1111/medu.14401

